# Locally calibrated error rates improve interpretability of AI scores and influence radiologist decision-making

**DOI:** 10.1101/2025.02.28.25323066

**Authors:** Maggie Chung, Micheal H. Bernstein, Lars J. Grimm, Adam Yala, Grayson L. Baird

**Affiliations:** Department of Radiology and Biomedical Imaging, University of California, San Francisco, CA. United States; Brown Radiology Human Factors Lab, Department of Radiology, The Warren Alpert Medical School, Brown University, and Brown University Health, Providence, RI. United States; Department of Radiology, Duke University, School of Medicine, Box 3808, DUMC, Durham, NC 27710; Computational Precision Health, University of California, Berkeley and University of California, San Francisco, CA. United States

**Author notes:** Corresponding Author. Maggie Chung, MD. Equal contribution (Maggie Chung, Micheal H. Bernstein).

## Abstract

**Introduction:** Artificial intelligence (AI) systems in radiology commonly generate case-level numeric scores intended to reflect the likelihood of underlying pathology. However, these scores are often difficult to interpret in clinical practice. We propose a framework for translating AI scores into clinically meaningful, locally calibrated error probabilities by providing the corresponding false discovery rate (FDR) and false omission rate (FOR) at each score threshold.

**Methods:** Using an open-source mammography AI model (Mirai), we estimated score-specific FDR and FOR across a range of thresholds using a retrospective cohort of 130,712 digital screening mammograms (907 positive, 129,805 negative). We then conducted a decision-making study to evaluate whether presenting FDR/FOR alongside AI scores influenced radiologist recall recommendations and confidence compared with AI scores alone.

**Results:** FDR and FOR varied substantially across AI score thresholds, ranging from 60.87% and 0.03%, respectively, at the low end of the score distribution to 99.26%% and 0.65% at the high end. In the decision-making study (n=21; 20 assessments per radiologist), recall increased with AI score in both conditions; however, recall was higher when AI scores were presented alone compared with scores accompanied by FDR/FOR (odds ratio 2.9, 95% CI [1.331, 6.417], p=0.0077). Confidence followed a U-shaped relationship with score and was higher when FDR/FOR were provided, particularly at intermediate scores.

**Conclusion:** Locally calibrated FDR and FOR provide a practical approach for translating AI scores into clinically interpretable probabilities. Presenting these measures alongside AI scores improves interpretability and is associated with changes in radiologist decision-making, supporting their use as a framework for clinical implementation of AI.

## Introduction

Artificial intelligence (AI) in medicine has expanded rapidly in recent years, particularly in radiology (1,2). While there has been considerable research on the integration of AI into clinical workflows, the interpretability and clinical transparency of AI outputs remain limited. In practice, AI systems commonly provide numeric output for each case, such as a risk or severity score (3–7). These scores are intended to reflect the likelihood of underlying pathology, however they have several limitations (8). First, AI scores are not readily interpretable, especially when they fall within the mid-range rather than at the extreme ends of the scale. For instance, if scores can range from 0.0 to 1.0, it is unclear what a score of 0.50 or 0.70 represents in a clinical context. This ambiguity may paradoxically increase a clinician’s uncertainty when evaluating a given case rather than providing useful guidance to inform a radiologist’s interpretation. Second, each AI algorithm has its own, often proprietary, scoring system (8). As a result, a score of 0.90 from one AI model may convey a different level of clinical concern than a score of 0.90 from another model, even for the same pathology. Moreover, different algorithms may use entirely different scoring scales, limiting direct comparisons between models. Finally, the relationship between AI score and underlying probability of pathology is not necessarily linear. For example, an increase in score from 0.20 to 0.30 may not correspond to the same change in risk as an increase from 0.60 to 0.70. Without a clear understanding of how scores relate to pathology, clinicians may struggle to determine what constitutes meaningful risk or a meaningful change in risk. This lack of calibration and interpretability reduces transparency, impedes shared decision-making, and limits trust in AI-assisted interpretations.

These concerns are particularly important in radiology, where decisions are often made under uncertainty and where the consequences of false-positive and false-negative interpretations are asymmetrical. Radiologists must often decide whether an equivocal finding should prompt intervention or follow-up. In this setting, AI has the potential to support decision-making, but only if its outputs can be interpreted in a way that is clinically meaningful. Raw scores alone do not provide this context. Traditional performance metrics such as sensitivity and specificity are commonly used for evaluating overall model performance, but they are less useful at the point of care because they are conditioned on the true disease state, which is unknown at the time of interpretation. Sensitivity refers to: “Of all the people who have the disease, what proportion will test positive?” Specificity refers to “Of all the people who do not have the disease, what proportion will test negative?” However, as noted above, at the time of decision making, the true disease status is unknown. Only the test result (AI score) is available to clinicians, and they typically want to know: “Given this AI score, what is the probability that the patient actually has (or does not have) the disease?”

To address this challenge, we propose pairing AI scores with the false discovery rate (FDR) and false omission rate (FOR) at each score threshold. FDR answers the clinically relevant question: among cases at or above a given AI score threshold, what proportion are negative for disease? FOR addresses the complementary question: among cases below that threshold, what proportion are positive for disease? Unlike raw AI scores, these quantities translate model output into error probabilities that are directly interpretable and relevant to clinical decision-making at the point of care. Importantly, they can be estimated using local historical data, allowing interpretation to reflect the patient population and disease prevalence of the practice in which the model is deployed. This is particularly important because predictive model performance and calibration are not fixed properties and likely vary substantially across populations and clinical environments (9). They can help create a more transparent and standardized language for interpreting AI output across systems that otherwise use different scoring scales (8,10). While FDR and FOR are established calibration-related metrics, they are generally used to characterize model performance at a population level rather than to support interpretation of specific AI score thresholds at the point of care. Here, we propose presenting score-specific, locally calibrated FDR and FOR values alongside AI outputs to provide clinically meaningful, decision-relevant context.

Whether this added context changes how radiologists use AI output remains uncertain. If locally calibrated FDR and FOR make AI scores more interpretable, they may influence both behavior and confidence, particularly for intermediate scores that are otherwise difficult to act on. In this study we first derived locally calibrated FDR and FOR across the range of scores from a mammography AI model using a large retrospective screening cohort. We then evaluated whether presenting radiologists with these error probabilities alongside AI scores altered recall decisions and confidence compared with presenting AI scores alone. We hypothesized that providing FDR and FOR would reduce recall relative to AI scores alone and increase radiologist confidence in decision-making.

## Methods

### Study design

This study comprised two components. First, we derived score-specific false discovery rates (FDR) and false omission rates (FOR) for a mammography artificial intelligence (AI) model using a large retrospective screening cohort. Second, we conducted a decision-making study to evaluate whether presenting these locally derived error probabilities alongside AI scores influenced radiologists’ recall recommendations and decision confidence, relative to presentation of AI scores alone. This HIPAA-compliant study was approved by the Institutional Review Board.

### Study sample

We conducted a retrospective review using a single-institution radiology database to identify 130,712 digital screening mammograms acquired between January 2006 and January 2023. All cases included at least one year of mammographic and/or clinical follow-up. Positive exams were defined as those with a histopathologic diagnosis of invasive breast carcinoma or ductal carcinoma in situ (DCIS) within 12 months of imaging. Negative exams were defined as those with at least 12 months of follow-up without a breast cancer diagnosis. Among the 129,805 cases, there were 907 positive exams.

### AI Model

To promote reproducibility and open science, we applied Mirai, an open-source AI model for mammogram-based risk prediction, to estimate 1-year breast cancer risk using 2D digital mammograms (11), though any AI system that generates an AI score could be used. No additional image processing was performed before the model application. The AI model was trained on a dataset independent of the one used for the present analyses.

### Derivation of score-specific FDR and FOR

Using the retrospective cohort, we estimated threshold-specific performance characteristics across the range of AI scores. For each score threshold, the FDR was defined as the proportion of examinations at or above the threshold that were cancer negative, and the FOR was defined as the proportion of examinations below the threshold that were cancer positive. These quantities were derived from local historical data to generate threshold-specific error probabilities calibrated to the screening population at our institution. Sensitivity, specificity, positive predictive value (PPV), and negative predictive value (NPV) were also calculated for descriptive purposes.

### Decision-making study

We conducted a decision-making study in which participants served as their own controls using Qualtrics (Provo, UT). Radiologists and fellows were first presented with 10 hypothetical screening mammography interpretation scenarios spanning 10 corresponding AI scores. Participants were told to assume a typical screening population with a breast cancer prevalence of approximately 5-6 cancers in every 1,000 mammograms, where the AI system had an ROC-AUC of 0.83, and AI scores could range from 0.00 to 1.00, with higher scores indicating greater likelihood of cancer, but the score was not an actual percentage.

Participants were then asked to imagine being on the fence about whether to give the case a BI-RADS 1/2 and recommend screening in one year or give it a BI-RADS 0 and recommend additional imaging, each accompanied by one of the 10 AI scores. The order of the AI scores was randomized. After a 4-week minimum washout period, participants were then asked to re-evaluate the cases, but this time each AI score was accompanied by its corresponding FDR/FOR values (see Table 1). For each scenario, participants indicated whether they would recommend recall for additional imaging evaluation and rated their confidence in that decision (from 0-100). Radiologists were recruited from three tertiary academic breast imaging centers XX, XX, XX). Vignettes are found in Supplemental Materials.

**Table 1.**
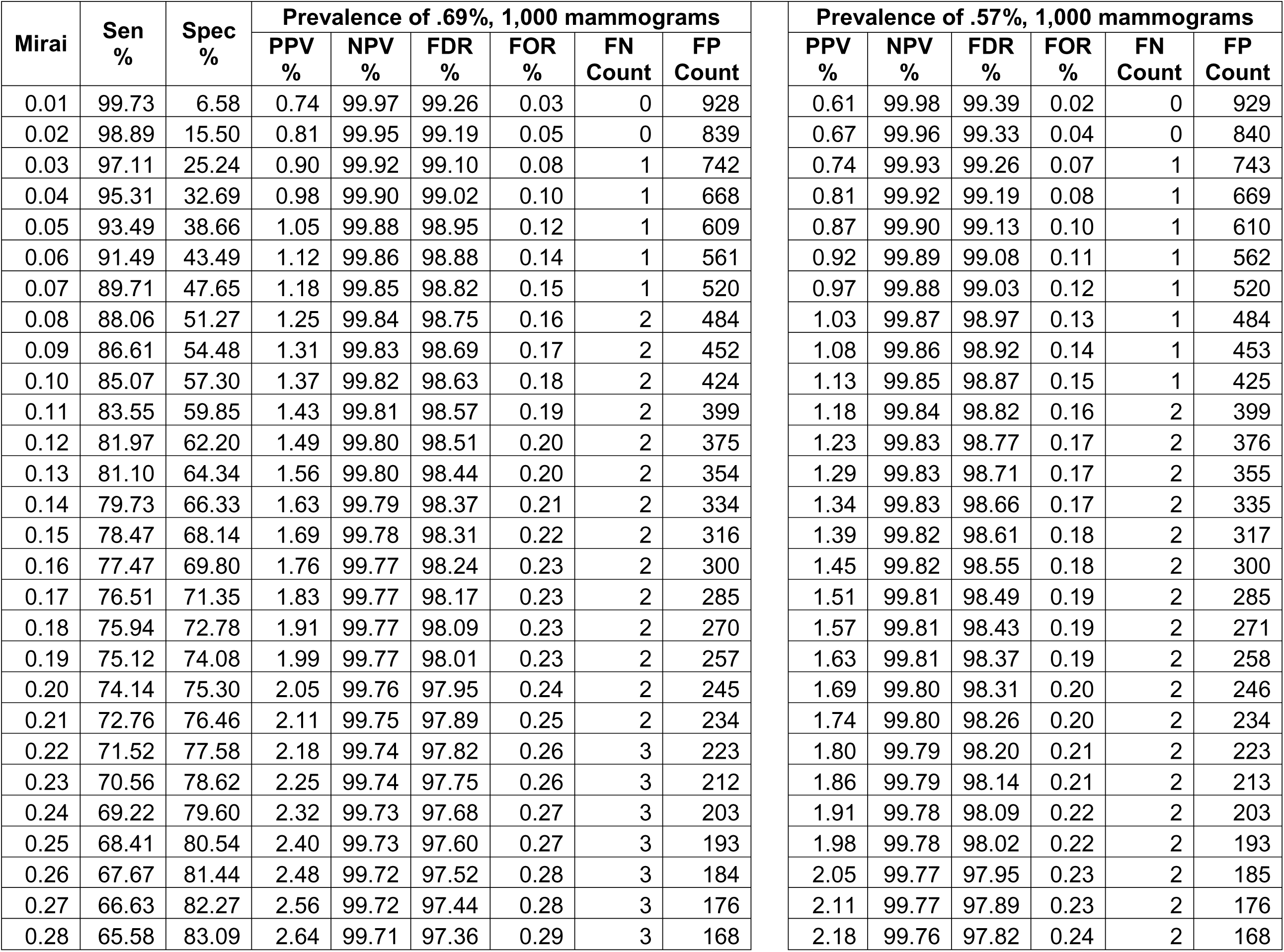
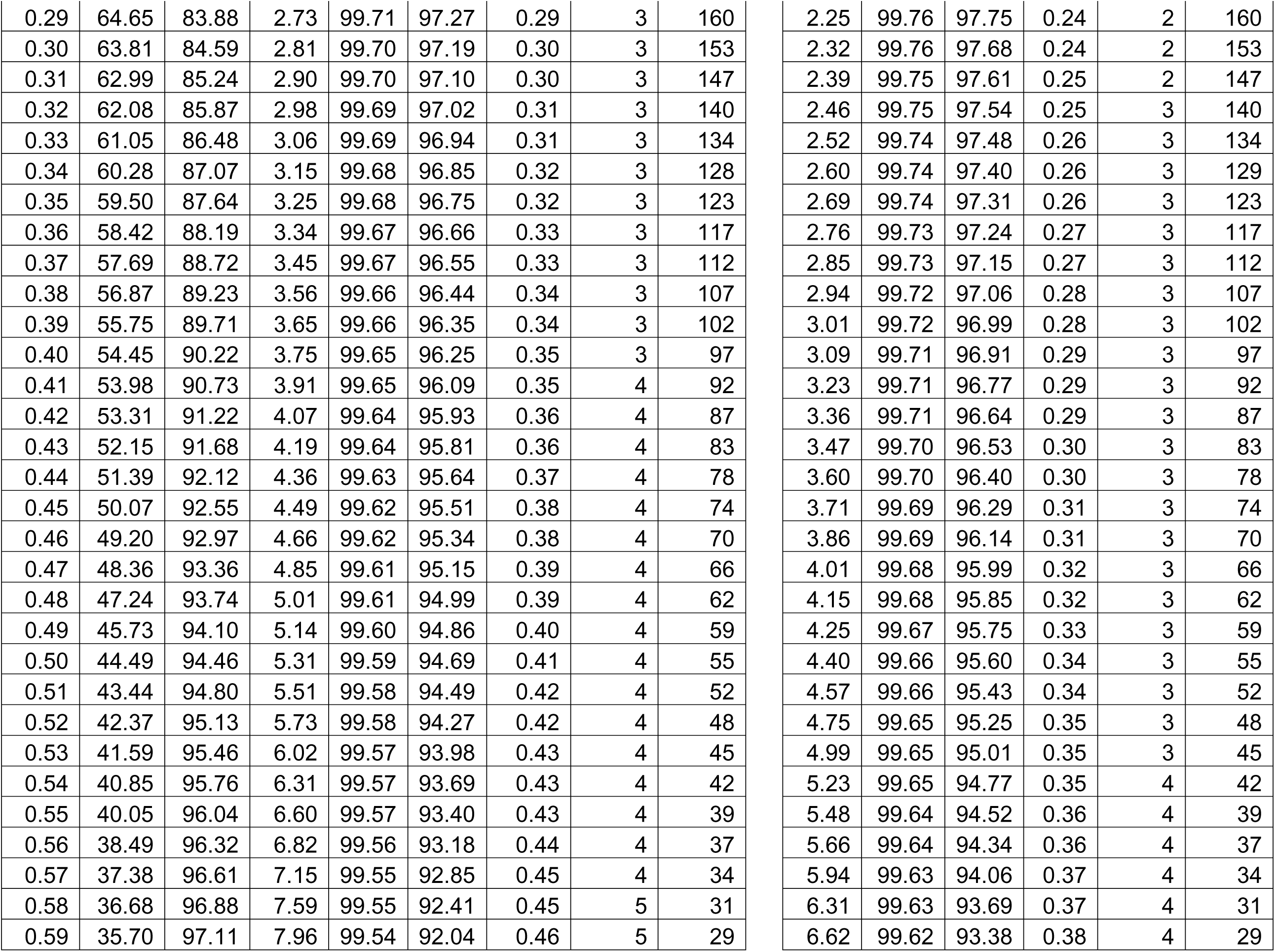
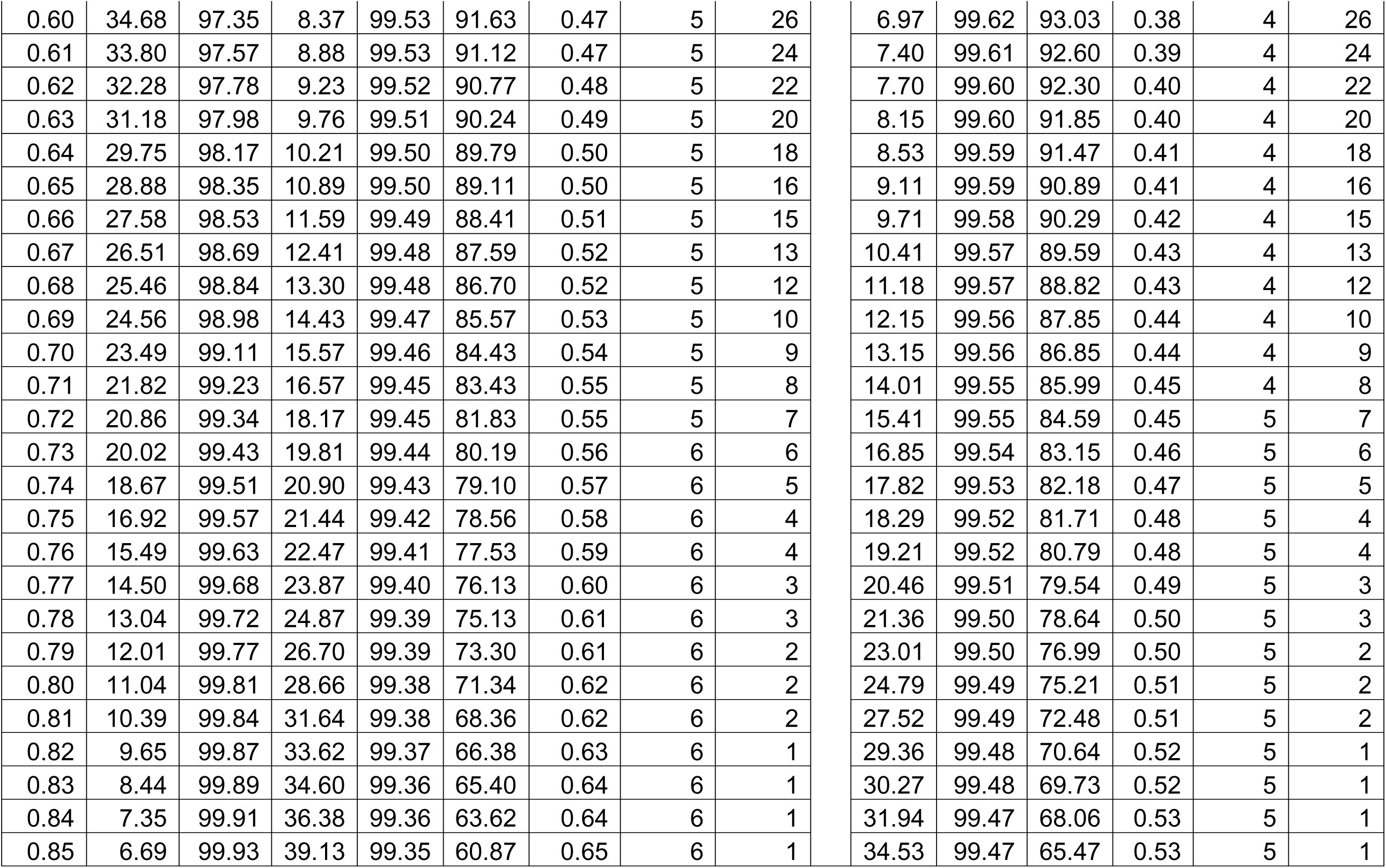
AI Score and other Metrics. Table Notes. AI scores ranging from 0.01 to 0.85 are provided along with the corresponding diagnostic performance metrics and outcomes (false positive and negative counts). Sen=Sensitivity, Spec=Specificity, PPV=Positive Predictive Value, FDR=False Discovery Rate (1-PPV), NPV=Negative Predictive Value, FOR=False Omission Rate (1-NPV), Prev.=prevalence, Rate (hypothetical number of mammograms read), #FN (of these 1,000, number of misses (false negatives)), #FP (of these 1,000, number of false alarms (false positives)).

| Mirai | Sen % | Spec % | Prevalence of .69%, 1,000 mammograms |  |  |  |  |  |
| --- | --- | --- | --- | --- | --- | --- | --- | --- |
|  |  |  | PPV % | NPV % | FDR % | FOR % | FN Count | FP Count |
| 0.01 | 99.73 | 6.58 | 0.74 | 99.97 | 99.26 | 0.03 | 0 | 928 |
| 0.02 | 98.89 | 15.50 | 0.81 | 99.95 | 99.19 | 0.05 | 0 | 839 |
| 0.03 | 97.11 | 25.24 | 0.90 | 99.92 | 99.10 | 0.08 | 1 | 742 |
| 0.04 | 95.31 | 32.69 | 0.98 | 99.90 | 99.02 | 0.10 | 1 | 668 |
| 0.05 | 93.49 | 38.66 | 1.05 | 99.88 | 98.95 | 0.12 | 1 | 609 |
| 0.06 | 91.49 | 43.49 | 1.12 | 99.86 | 98.88 | 0.14 | 1 | 561 |
| 0.07 | 89.71 | 47.65 | 1.18 | 99.85 | 98.82 | 0.15 | 1 | 520 |
| 0.08 | 88.06 | 51.27 | 1.25 | 99.84 | 98.75 | 0.16 | 2 | 484 |
| 0.09 | 86.61 | 54.48 | 1.31 | 99.83 | 98.69 | 0.17 | 2 | 452 |
| 0.10 | 85.07 | 57.30 | 1.37 | 99.82 | 98.63 | 0.18 | 2 | 424 |
| 0.11 | 83.55 | 59.85 | 1.43 | 99.81 | 98.57 | 0.19 | 2 | 399 |
| 0.12 | 81.97 | 62.20 | 1.49 | 99.80 | 98.51 | 0.20 | 2 | 375 |
| 0.13 | 81.10 | 64.34 | 1.56 | 99.80 | 98.44 | 0.20 | 2 | 354 |
| 0.14 | 79.73 | 66.33 | 1.63 | 99.79 | 98.37 | 0.21 | 2 | 334 |
| 0.15 | 78.47 | 68.14 | 1.69 | 99.78 | 98.31 | 0.22 | 2 | 316 |
| 0.16 | 77.47 | 69.80 | 1.76 | 99.77 | 98.24 | 0.23 | 2 | 300 |
| 0.17 | 76.51 | 71.35 | 1.83 | 99.77 | 98.17 | 0.23 | 2 | 285 |
| 0.18 | 75.94 | 72.78 | 1.91 | 99.77 | 98.09 | 0.23 | 2 | 270 |
| 0.19 | 75.12 | 74.08 | 1.99 | 99.77 | 98.01 | 0.23 | 2 | 257 |
| 0.20 | 74.14 | 75.30 | 2.05 | 99.76 | 97.95 | 0.24 | 2 | 245 |
| 0.21 | 72.76 | 76.46 | 2.11 | 99.75 | 97.89 | 0.25 | 2 | 234 |
| 0.22 | 71.52 | 77.58 | 2.18 | 99.74 | 97.82 | 0.26 | 3 | 223 |
| 0.23 | 70.56 | 78.62 | 2.25 | 99.74 | 97.75 | 0.26 | 3 | 212 |
| 0.24 | 69.22 | 79.60 | 2.32 | 99.73 | 97.68 | 0.27 | 3 | 203 |
| 0.25 | 68.41 | 80.54 | 2.40 | 99.73 | 97.60 | 0.27 | 3 | 193 |
| 0.26 | 67.67 | 81.44 | 2.48 | 99.72 | 97.52 | 0.28 | 3 | 184 |
| 0.27 | 66.63 | 82.27 | 2.56 | 99.72 | 97.44 | 0.28 | 3 | 176 |
| 0.28 | 65.58 | 83.09 | 2.64 | 99.71 | 97.36 | 0.29 | 3 | 168 |

| Prevalence of .57%, 1,000 mammograms |  |  |  |  |  |
| --- | --- | --- | --- | --- | --- |
| PPV % | NPV % | FDR % | FOR % | FN Count | FP Count |
| 0.61 | 99.98 | 99.39 | 0.02 | 0 | 929 |
| 0.67 | 99.96 | 99.33 | 0.04 | 0 | 840 |
| 0.74 | 99.93 | 99.26 | 0.07 | 1 | 743 |
| 0.81 | 99.92 | 99.19 | 0.08 | 1 | 669 |
| 0.87 | 99.90 | 99.13 | 0.10 | 1 | 610 |
| 0.92 | 99.89 | 99.08 | 0.11 | 1 | 562 |
| 0.97 | 99.88 | 99.03 | 0.12 | 1 | 520 |
| 1.03 | 99.87 | 98.97 | 0.13 | 1 | 484 |
| 1.08 | 99.86 | 98.92 | 0.14 | 1 | 453 |
| 1.13 | 99.85 | 98.87 | 0.15 | 1 | 425 |
| 1.18 | 99.84 | 98.82 | 0.16 | 2 | 399 |
| 1.23 | 99.83 | 98.77 | 0.17 | 2 | 376 |
| 1.29 | 99.83 | 98.71 | 0.17 | 2 | 355 |
| 1.34 | 99.83 | 98.66 | 0.17 | 2 | 335 |
| 1.39 | 99.82 | 98.61 | 0.18 | 2 | 317 |
| 1.45 | 99.82 | 98.55 | 0.18 | 2 | 300 |
| 1.51 | 99.81 | 98.49 | 0.19 | 2 | 285 |
| 1.57 | 99.81 | 98.43 | 0.19 | 2 | 271 |
| 1.63 | 99.81 | 98.37 | 0.19 | 2 | 258 |
| 1.69 | 99.80 | 98.31 | 0.20 | 2 | 246 |
| 1.74 | 99.80 | 98.26 | 0.20 | 2 | 234 |
| 1.80 | 99.79 | 98.20 | 0.21 | 2 | 223 |
| 1.86 | 99.79 | 98.14 | 0.21 | 2 | 213 |
| 1.91 | 99.78 | 98.09 | 0.22 | 2 | 203 |
| 1.98 | 99.78 | 98.02 | 0.22 | 2 | 193 |
| 2.05 | 99.77 | 97.95 | 0.23 | 2 | 185 |
| 2.11 | 99.77 | 97.89 | 0.23 | 2 | 176 |
| 2.18 | 99.76 | 97.82 | 0.24 | 2 | 168 |
| 0.29 | 64.65 | 83.88 | 2.73 | 99.71 | 97.27 | 0.29 | 3 | 160 |
| 0.30 | 63.81 | 84.59 | 2.81 | 99.70 | 97.19 | 0.30 | 3 | 153 |
| 0.31 | 62.99 | 85.24 | 2.90 | 99.70 | 97.10 | 0.30 | 3 | 147 |
| 0.32 | 62.08 | 85.87 | 2.98 | 99.69 | 97.02 | 0.31 | 3 | 140 |
| 0.33 | 61.05 | 86.48 | 3.06 | 99.69 | 96.94 | 0.31 | 3 | 134 |
| 0.34 | 60.28 | 87.07 | 3.15 | 99.68 | 96.85 | 0.32 | 3 | 128 |
| 0.35 | 59.50 | 87.64 | 3.25 | 99.68 | 96.75 | 0.32 | 3 | 123 |
| 0.36 | 58.42 | 88.19 | 3.34 | 99.67 | 96.66 | 0.33 | 3 | 117 |
| 0.37 | 57.69 | 88.72 | 3.45 | 99.67 | 96.55 | 0.33 | 3 | 112 |
| 0.38 | 56.87 | 89.23 | 3.56 | 99.66 | 96.44 | 0.34 | 3 | 107 |
| 0.39 | 55.75 | 89.71 | 3.65 | 99.66 | 96.35 | 0.34 | 3 | 102 |
| 0.40 | 54.45 | 90.22 | 3.75 | 99.65 | 96.25 | 0.35 | 3 | 97 |
| 0.41 | 53.98 | 90.73 | 3.91 | 99.65 | 96.09 | 0.35 | 4 | 92 |
| 0.42 | 53.31 | 91.22 | 4.07 | 99.64 | 95.93 | 0.36 | 4 | 87 |
| 0.43 | 52.15 | 91.68 | 4.19 | 99.64 | 95.81 | 0.36 | 4 | 83 |
| 0.44 | 51.39 | 92.12 | 4.36 | 99.63 | 95.64 | 0.37 | 4 | 78 |
| 0.45 | 50.07 | 92.55 | 4.49 | 99.62 | 95.51 | 0.38 | 4 | 74 |
| 0.46 | 49.20 | 92.97 | 4.66 | 99.62 | 95.34 | 0.38 | 4 | 70 |
| 0.47 | 48.36 | 93.36 | 4.85 | 99.61 | 95.15 | 0.39 | 4 | 66 |
| 0.48 | 47.24 | 93.74 | 5.01 | 99.61 | 94.99 | 0.39 | 4 | 62 |
| 0.49 | 45.73 | 94.10 | 5.14 | 99.60 | 94.86 | 0.40 | 4 | 59 |
| 0.50 | 44.49 | 94.46 | 5.31 | 99.59 | 94.69 | 0.41 | 4 | 55 |
| 0.51 | 43.44 | 94.80 | 5.51 | 99.58 | 94.49 | 0.42 | 4 | 52 |
| 0.52 | 42.37 | 95.13 | 5.73 | 99.58 | 94.27 | 0.42 | 4 | 48 |
| 0.53 | 41.59 | 95.46 | 6.02 | 99.57 | 93.98 | 0.43 | 4 | 45 |
| 0.54 | 40.85 | 95.76 | 6.31 | 99.57 | 93.69 | 0.43 | 4 | 42 |
| 0.55 | 40.05 | 96.04 | 6.60 | 99.57 | 93.40 | 0.43 | 4 | 39 |
| 0.56 | 38.49 | 96.32 | 6.82 | 99.56 | 93.18 | 0.44 | 4 | 37 |
| 0.57 | 37.38 | 96.61 | 7.15 | 99.55 | 92.85 | 0.45 | 4 | 34 |
| 0.58 | 36.68 | 96.88 | 7.59 | 99.55 | 92.41 | 0.45 | 5 | 31 |
| 0.59 | 35.70 | 97.11 | 7.96 | 99.54 | 92.04 | 0.46 | 5 | 29 |

|  |  |  |  |  |  |
| --- | --- | --- | --- | --- | --- |
| 2.25 | 99.76 | 97.75 | 0.24 | 2 | 160 |
| 2.32 | 99.76 | 97.68 | 0.24 | 2 | 153 |
| 2.39 | 99.75 | 97.61 | 0.25 | 2 | 147 |
| 2.46 | 99.75 | 97.54 | 0.25 | 3 | 140 |
| 2.52 | 99.74 | 97.48 | 0.26 | 3 | 134 |
| 2.60 | 99.74 | 97.40 | 0.26 | 3 | 129 |
| 2.69 | 99.74 | 97.31 | 0.26 | 3 | 123 |
| 2.76 | 99.73 | 97.24 | 0.27 | 3 | 117 |
| 2.85 | 99.73 | 97.15 | 0.27 | 3 | 112 |
| 2.94 | 99.72 | 97.06 | 0.28 | 3 | 107 |
| 3.01 | 99.72 | 96.99 | 0.28 | 3 | 102 |
| 3.09 | 99.71 | 96.91 | 0.29 | 3 | 97 |
| 3.23 | 99.71 | 96.77 | 0.29 | 3 | 92 |
| 3.36 | 99.71 | 96.64 | 0.29 | 3 | 87 |
| 3.47 | 99.70 | 96.53 | 0.30 | 3 | 83 |
| 3.60 | 99.70 | 96.40 | 0.30 | 3 | 78 |
| 3.71 | 99.69 | 96.29 | 0.31 | 3 | 74 |
| 3.86 | 99.69 | 96.14 | 0.31 | 3 | 70 |
| 4.01 | 99.68 | 95.99 | 0.32 | 3 | 66 |
| 4.15 | 99.68 | 95.85 | 0.32 | 3 | 62 |
| 4.25 | 99.67 | 95.75 | 0.33 | 3 | 59 |
| 4.40 | 99.66 | 95.60 | 0.34 | 3 | 55 |
| 4.57 | 99.66 | 95.43 | 0.34 | 3 | 52 |
| 4.75 | 99.65 | 95.25 | 0.35 | 3 | 48 |
| 4.99 | 99.65 | 95.01 | 0.35 | 3 | 45 |
| 5.23 | 99.65 | 94.77 | 0.35 | 4 | 42 |
| 5.48 | 99.64 | 94.52 | 0.36 | 4 | 39 |
| 5.66 | 99.64 | 94.34 | 0.36 | 4 | 37 |
| 5.94 | 99.63 | 94.06 | 0.37 | 4 | 34 |
| 6.31 | 99.63 | 93.69 | 0.37 | 4 | 31 |
| 6.62 | 99.62 | 93.38 | 0.38 | 4 | 29 |

**Table 1. AI Score and other Metrics.**
|  |  |  |  |  |  |  |  |  |  |  |  |  |  |  |
| --- | --- | --- | --- | --- | --- | --- | --- | --- | --- | --- | --- | --- | --- | --- |
| 0.60 | 34.68 | 97.35 | 8.37 | 99.53 | 91.63 | 0.47 | 5 | 26 | 6.97 | 99.62 | 93.03 | 0.38 | 4 | 26 |
| 0.61 | 33.80 | 97.57 | 8.88 | 99.53 | 91.12 | 0.47 | 5 | 24 | 7.40 | 99.61 | 92.60 | 0.39 | 4 | 24 |
| 0.62 | 32.28 | 97.78 | 9.23 | 99.52 | 90.77 | 0.48 | 5 | 22 | 7.70 | 99.60 | 92.30 | 0.40 | 4 | 22 |
| 0.63 | 31.18 | 97.98 | 9.76 | 99.51 | 90.24 | 0.49 | 5 | 20 | 8.15 | 99.60 | 91.85 | 0.40 | 4 | 20 |
| 0.64 | 29.75 | 98.17 | 10.21 | 99.50 | 89.79 | 0.50 | 5 | 18 | 8.53 | 99.59 | 91.47 | 0.41 | 4 | 18 |
| 0.65 | 28.88 | 98.35 | 10.89 | 99.50 | 89.11 | 0.50 | 5 | 16 | 9.11 | 99.59 | 90.89 | 0.41 | 4 | 16 |
| 0.66 | 27.58 | 98.53 | 11.59 | 99.49 | 88.41 | 0.51 | 5 | 15 | 9.71 | 99.58 | 90.29 | 0.42 | 4 | 15 |
| 0.67 | 26.51 | 98.69 | 12.41 | 99.48 | 87.59 | 0.52 | 5 | 13 | 10.41 | 99.57 | 89.59 | 0.43 | 4 | 13 |
| 0.68 | 25.46 | 98.84 | 13.30 | 99.48 | 86.70 | 0.52 | 5 | 12 | 11.18 | 99.57 | 88.82 | 0.43 | 4 | 12 |
| 0.69 | 24.56 | 98.98 | 14.43 | 99.47 | 85.57 | 0.53 | 5 | 10 | 12.15 | 99.56 | 87.85 | 0.44 | 4 | 10 |
| 0.70 | 23.49 | 99.11 | 15.57 | 99.46 | 84.43 | 0.54 | 5 | 9 | 13.15 | 99.56 | 86.85 | 0.44 | 4 | 9 |
| 0.71 | 21.82 | 99.23 | 16.57 | 99.45 | 83.43 | 0.55 | 5 | 8 | 14.01 | 99.55 | 85.99 | 0.45 | 4 | 8 |
| 0.72 | 20.86 | 99.34 | 18.17 | 99.45 | 81.83 | 0.55 | 5 | 7 | 15.41 | 99.55 | 84.59 | 0.45 | 5 | 7 |
| 0.73 | 20.02 | 99.43 | 19.81 | 99.44 | 80.19 | 0.56 | 6 | 6 | 16.85 | 99.54 | 83.15 | 0.46 | 5 | 6 |
| 0.74 | 18.67 | 99.51 | 20.90 | 99.43 | 79.10 | 0.57 | 6 | 5 | 17.82 | 99.53 | 82.18 | 0.47 | 5 | 5 |
| 0.75 | 16.92 | 99.57 | 21.44 | 99.42 | 78.56 | 0.58 | 6 | 4 | 18.29 | 99.52 | 81.71 | 0.48 | 5 | 4 |
| 0.76 | 15.49 | 99.63 | 22.47 | 99.41 | 77.53 | 0.59 | 6 | 4 | 19.21 | 99.52 | 80.79 | 0.48 | 5 | 4 |
| 0.77 | 14.50 | 99.68 | 23.87 | 99.40 | 76.13 | 0.60 | 6 | 3 | 20.46 | 99.51 | 79.54 | 0.49 | 5 | 3 |
| 0.78 | 13.04 | 99.72 | 24.87 | 99.39 | 75.13 | 0.61 | 6 | 3 | 21.36 | 99.50 | 78.64 | 0.50 | 5 | 3 |
| 0.79 | 12.01 | 99.77 | 26.70 | 99.39 | 73.30 | 0.61 | 6 | 2 | 23.01 | 99.50 | 76.99 | 0.50 | 5 | 2 |
| 0.80 | 11.04 | 99.81 | 28.66 | 99.38 | 71.34 | 0.62 | 6 | 2 | 24.79 | 99.49 | 75.21 | 0.51 | 5 | 2 |
| 0.81 | 10.39 | 99.84 | 31.64 | 99.38 | 68.36 | 0.62 | 6 | 2 | 27.52 | 99.49 | 72.48 | 0.51 | 5 | 2 |
| 0.82 | 9.65 | 99.87 | 33.62 | 99.37 | 66.38 | 0.63 | 6 | 1 | 29.36 | 99.48 | 70.64 | 0.52 | 5 | 1 |
| 0.83 | 8.44 | 99.89 | 34.60 | 99.36 | 65.40 | 0.64 | 6 | 1 | 30.27 | 99.48 | 69.73 | 0.52 | 5 | 1 |
| 0.84 | 7.35 | 99.91 | 36.38 | 99.36 | 63.62 | 0.64 | 6 | 1 | 31.94 | 99.47 | 68.06 | 0.53 | 5 | 1 |
| 0.85 | 6.69 | 99.93 | 39.13 | 99.35 | 60.87 | 0.65 | 6 | 1 | 34.53 | 99.47 | 65.47 | 0.53 | 5 | 1 |

### Statistical analysis

All analyses were conducted using SAS 9.4 (SAS Cary, NC). For the local calibration study, sensitivity, specificity, PPV, NPV (and their complements), were estimated using the LOGISTIC procedure with the %ROCPLOT macro. The base rate of cancer was 0.69%. We also use a base of 0.57% as an example when local historical data are unavailable. For the decision-making study, recall rates were examined using generalized linear mixed modeling (GLMM) with experimental condition (AI scores with vs. without FDR/FOR) and AI score (0.05–0.85) entered as main effects and their interaction. Models assumed a binary distribution with logit link, random intercepts and slopes, and a compound symmetry covariance structure. To evaluate the relationship between confidence and AI score, GLMM incorporating natural cubic spline terms with three equally spaced knots were fit by AI Score. Inter-rater agreement was evaluated using Fleiss’ Kappa. Alpha was established a priori at 0.05, and all interval estimates were calculated for 95% confidence.

## Results

### Score-specific local calibration of AI output

As demonstrated in Table 1 and Figures 1 and 2, we examined FDR and FOR values across the full range of AI score thresholds. For brevity, these values are provided at 0.01-unit increments from 0.01 to 0.85 (inclusive). Also included in Table 1, for reference, are sensitivity, specificity, NPV, PPV, and prevalence. Across the retrospective screening cohort, FDR and FOR varied substantially across the range of AI score thresholds, demonstrating that equivalent numeric changes in raw AI score did not necessarily correspond to uniform changes in error probabilities. At the lower end of the score distribution, FDR demonstrates a slow initial decline followed by a pronounced, elbow-like drop. Conversely, the FOR increases rapidly at lower scores and then transitions to a more gradual, nearly constant rate of increase. Using the study cohort estimates, FDR ranged from 99.26% at a threshold of 0.01 to 60.87% at a threshold of 0.85, whereas FOR ranged from 0.03% to 0.65% across the same threshold range. At an AI score threshold of 0.50, the FDR was 94.69% and the FOR was 0.41%, indicating that among examinations with scores at or above 0.50, approximately 95% were cancer negative, whereas among examinations below 0.50, approximately 0.4% were cancer positive. At a threshold of 0.70, the FDR decreased to 84.43% and the FOR increased to 0.54%.

**Figure 1.**
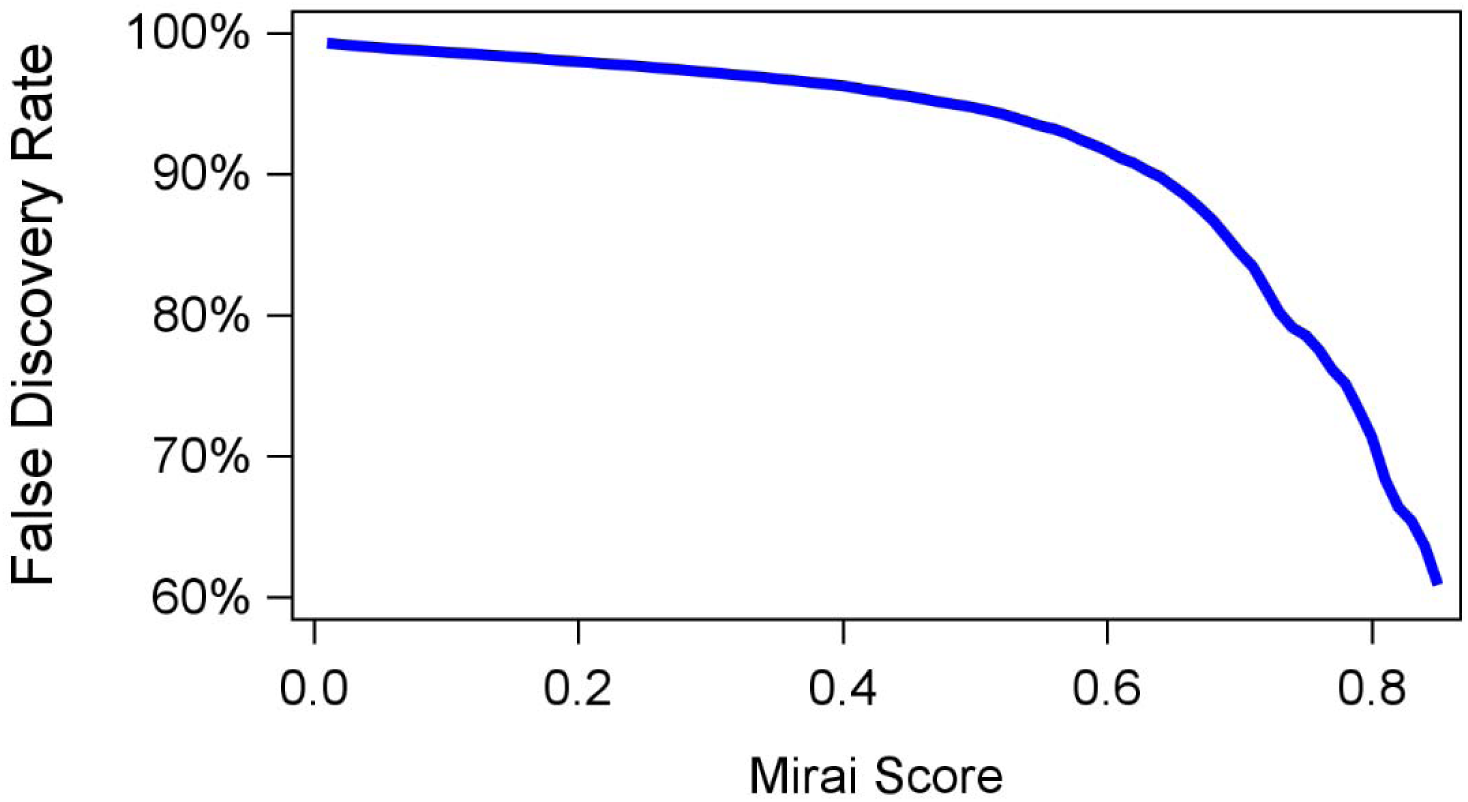
Relationship between Mirai Scores and False Discovery Rate. X-Axis is Mirai Score (0.05 to 0.85) and Y-axis is false discovery rate (60% to 100%). Blue line visualizes the FDR value at each Mirai Score.

**Figure 2.**
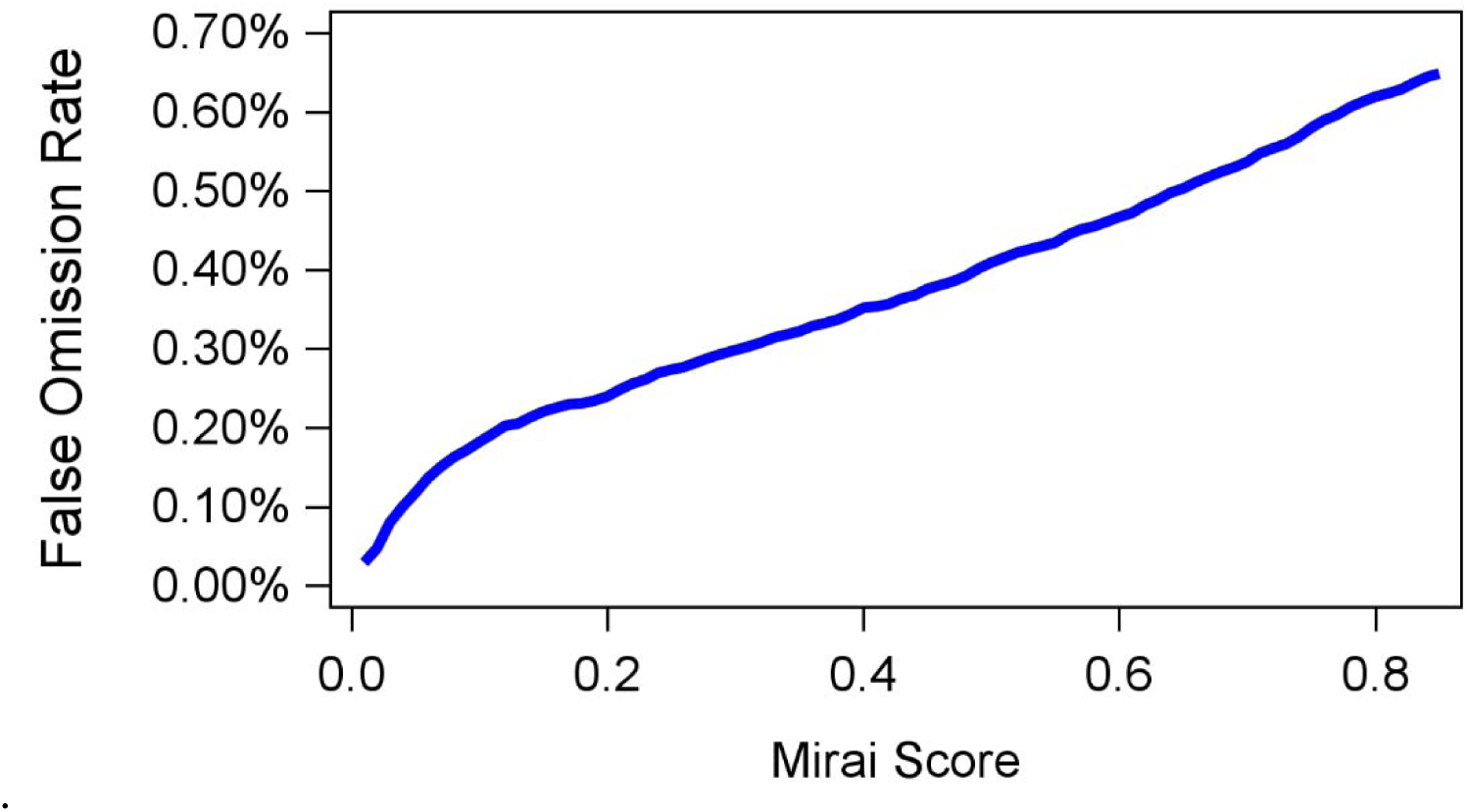
Relationship between Mirai Scores and False Omission Rate. X-Axis is Mirai Score (0.05 to 0.85) and Y-axis is false omission rate (00.0% to 0.60%). Blue line visualizes the FOR value at each Mirai Score.

The clinical implications of a change in AI score also varied across the score range. For example, increasing the score threshold from 0.01 to 0.10 was associated with a decrease in FDR from 99.26% to 98.63% (0.62 absolute delta), and an increase in FOR, from 0.03% to 0.18% (.15 absolute delta). In contrast, increasing the threshold from 0.70 to 0.80 was associated with a decrease in FDR from 84.43% to 71.34% (13.09 absolute delta), whereas FOR changed from 0.54% to .62% (.08 absolute change). Thus, identical absolute changes in AI score can correspond to substantially different relative and absolute changes in error probabilities.

### Decision-making Study

A total of 21 participants completed the study, including 14 fellowship-trained breast imaging radiologists and 7 current breast imaging fellows. Sixteen participants were women (76.2%) and five were men (23.8%). Among attending radiologists, years of post-training experience included 1–5 years (n=8), 6–10 years (n=3), and >10 years (n=3). Each participant provided 20 assessments (10 pre and post). Sensitivity analyses excluding fellows yielded similar findings.

### Impact of Providing FDR/FOR on Recall Recommendations

As illustrated in Figure 3, the slopes describing the relationship between AI scores and recall rates failed to be different from each other, p=0.9595 (interaction effect): as AI score increased, radiologist recall increased for both, p<0.0001 (main effect). However, radiologists’ recall rate was higher when the AI scores were provided alone compared with the same AI scores accompanied with their corresponding FDR/FOR—an almost 3-fold odds increase (OR: 2.923, 95% CI [1.331, 6.417], p=0.0077, main effect), though at the upper levels of the AI score, this difference diminished.

**Figure 3.**
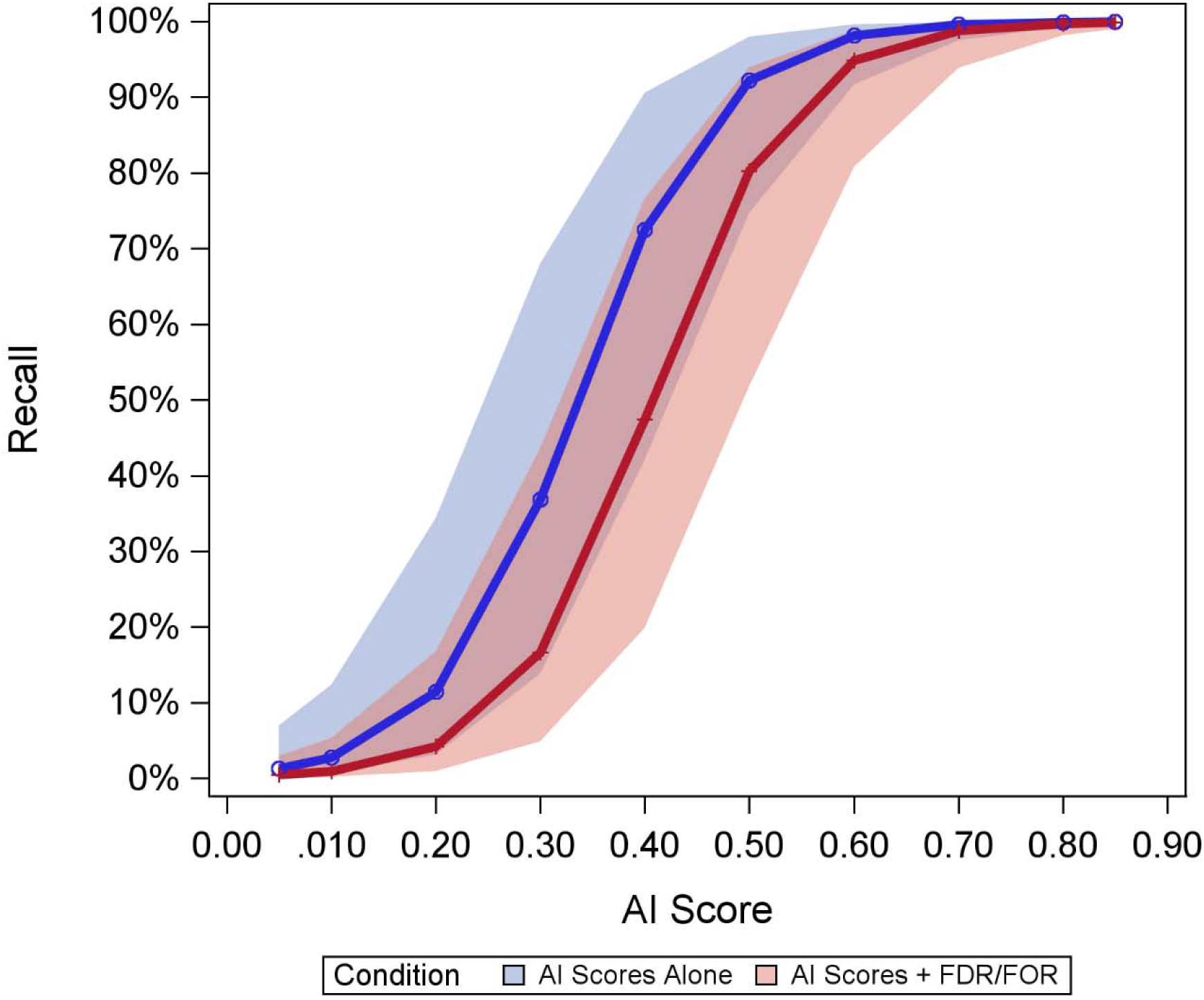
Percentage of radiologists who would recall as a function of AI score and condition (AI scores alone, blue; AI scores + FDR/FOR). X-Axis is the AI Score (0.05 to 0.85) and Y-axis is recall rate (0% to 100%).

### Impact of Providing FDR/FOR on Radiologist Confidence

As illustrated in Figure 4, reported confidence followed an approximant U-shape curve, revealing greater confidence in the extremes of AI scores and diminishing in the middle (main effect), p<0.0001. Confidence was higher for higher AI scores, likely due to being confident in recalling, while confidence was relatively lower for lower AI scores, as confidence likely reflected not recalling, which introduces risk. Reported confidence by radiologists was higher when radiologists were provided FDR/FOR with the AI scores compared to when they were provided the AI scores only, but this was true for AI scores under 0.60; as the AI scores increased, confidence by condition converged, with a significant interaction effect (p=0.0009).

**Figure 4.**
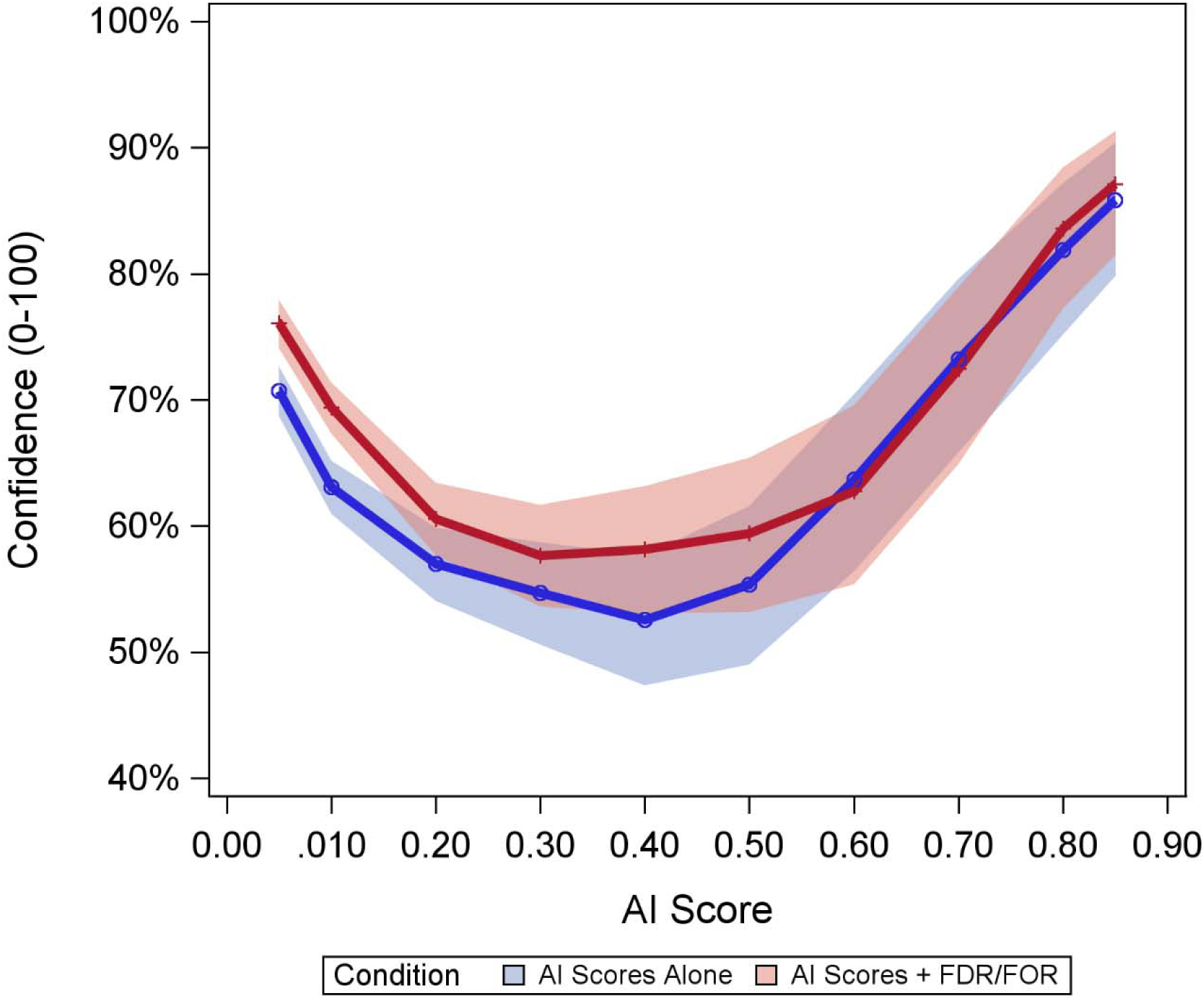
Radiologist confidence as a function of AI score and condition (AI scores alone, blue; AI scores + FDR/FOR). X-Axis is the AI Score (0.05 to 0.85), and Y-axis is confidence (0% to 100%)

### Impact of Inter-Rater Agreement

The Fleiss’ Kappa was moderate with AI scores alone (0.44, SE: 0.024) and AI scores with FDR/FOR (0.49, SE: 0.024), p=0.0104.

## Discussion

In this study, we demonstrate and evaluated a practical framework for translating AI scores into clinically interpretable, locally calibrated error probabilities. Although FDR decreased and FOR increased monotonically across increasing AI score thresholds, the nonlinear nature of these relationships demonstrate that raw AI scores do not provide an intuitive or directly interpretable estimate of clinically meaningful probability. In our decision-making experiment, we found that presenting radiologists with score-specific FDR and FOR, rather than the AI score alone, was associated with lower recall rates and greater decision confidence, particularly in the intermediate score range where AI scores are often most difficult to interpret and their clinical implications may be uncertain. Together, these findings suggest that score-specific FDR and FOR may improve the interpretability of AI output and support more informed decision-making.

By reporting the error rates (FDR and FOR) corresponding to each score, scores can now be contextualized using a common language—the language of probability of error. While traditional metrics such as sensitivity and specificity are critical in the assessment of overall model performance, they have limited utility for individual cases because they are conditional on the true disease state. Critically, however, this information is *unknown when a clinician is reviewing a case* – indeed, true disease state is the very thing a radiologist tries to determine and is ipso facto unknown *a priori*. In contrast, FDR and FOR are estimates of the proportion of FNs and FPs (respectively) conditional on the AI score, *and the score is the only piece of information available to clinicians at the time of decision-making*. Thus, FDR and FOR, unlike sensitivity and specificity, are predicated on the only information about a case which a radiologist can know at the time of reviewing the case.

Our results also show that the same absolute change in AI score may correspond to very different changes in FDR and FOR depending on the threshold considered. This is important because raw scores may otherwise appear more directly interpretable than they are in practice. Furthermore, a score of 0.50 on a 0–1.00 scale may be misinterpreted as having inherent probabilistic meaning, despite the absence of direct calibration between the raw score and clinical probability. Indeed, psychological research suggests that individuals base judgments on subjective perceptions of probability rather than objective probability itself (12). As such, presenting threshold-specific error rates can help to correct potentially false beliefs by providing a clearer representation of the probability and tradeoff between false-positive and false-negative decisions across the score range.

In the decision-making study, we found that this additional context influences radiologist behavior. When shown AI scores alone, radiologists were more likely to recommend recall than when shown the same scores with corresponding FDR and FOR. One possible explanation is that, in the absence of calibration context, radiologists may interpret a raw score as more concerning than its local error rates would support. Notably, radiologist confidence was lowest in the middle of the score range and improved when FDR and FOR were shown. This suggests that providing FDR and FOR may be most useful where AI scores are more ambiguous and difficult to interpret.

A notable strength of our proposed approach is that FDR and FOR are locally derived. The clinical meaning of an AI score depends in part on disease prevalence and case mix in the population in which the model is deployed. Vendor-defined thresholds may not fully reflect these local conditions. Estimating FDR and FOR from site-specific historical data provides context that is directly relevant to local practice. This framework may also facilitate comparisons across AI systems and model versions. Different models may use different proprietary scoring scales, and similar raw scores may correspond to different error profiles. Expressing model output in terms of FDR and FOR, however, provides a common probabilistic framework for interpreting thresholds across systems and, potentially, across successive model updates. In other words, while the meaning of scores across different systems and models may differ, the meaning of error rates are always the same.

In addition to their relevance for radiologist decision-making, these error rates may also be important for patients, as recent research indicates (13). As more patients have access to their exams and associated AI outputs, there may be a greater need for outputs that can be understood beyond the clinician alone. Threshold-specific error rates may provide a more interpretable way to communicate the meaning of AI output and its associated uncertainty.

This study has several limitations. First, the FDR and FOR estimates were derived from a single-institution retrospective cohort and reflect the local patient population and disease prevalence. These estimates are intended to illustrate a framework in which FDR and FOR can be calculated from local historical data. Second, the decision-making study was vignette-based rather than conducted in a prospective clinical setting. Images were not included in order to isolate the effect of AI score presentation and minimize confounding variables. Although this design allowed for controlled comparison of presentation formats, it may not fully reflect clinical setting. Prospective evaluation will be needed to assess the impact of this approach on clinical decision-making. Third, the Mirai AI model was used as an illustrative example to demonstrate how FDR and FOR can be derived for an AI system using historical data. Yet, the purpose of the retrospective study was not to evaluate the performance of this specific model. The proposed framework is intended to be generalizable to other AI systems and clinical applications. Fourth, reliable estimation of threshold-specific FDR and FOR requires sufficiently large datasets, particularly for low-prevalence outcomes such breast cancer in a screening setting. Smaller datasets may yield unstable estimates, especially at extreme score thresholds. Finally, FDR and FOR estimates may change over time as underlying data distributions evolve due to shifts in patient populations or disease prevalence. Periodic recalibration may be necessary.

In summary, AI scores alone may be difficult to interpret in clinical practice. Locally calibrated FDR and FOR provide a practical way to translate AI scores into clinically meaningful probabilities. In this study, presenting these measures alongside AI scores was associated with lower recall rates and greater confidence, particularly in the intermediate score range. These findings support the use of threshold-specific local error rates as a practical strategy to improve the interpretability of AI output in clinical practice.

## Data Availability

Data generated or analyzed during the study are available upon reasonable request to the corresponding author.

## Author Contributions

MC contributed to the conceptualization, methodology, formal analysis, investigation of the study, and writing of the manuscript. LJG contributed to the conceptualization, methodology, and investigation of the study, and writing of the manuscript. MHB contributed to the conceptualization, methodology, formal analysis, and investigation of the study, and writing of the manuscript. AY contributed to the conceptualization and writing of the manuscript. GLB contributed to the conceptualization, methodology, formal analysis, and investigation of the study, and writing of the manuscript. All authors read and approved the final manuscript.

## Abbreviations

AI: Artificial intelligence
FDR: False discovery rate
FOR: Fasel omission rate
PPV: Positive predictive value
NPV: Negative predictive value
DCIS: ductal carcinoma in situ

## Supplemental Materials

### Vignette Sessions

#### Introduction

This study asks you to visualize **one specific clinical scenario** where you are reading a screening mammogram and are undecided about whether to recommend a recall for additional imaging. **Essentially, you are on the fence about whether to give the case a BI-RADS: 1/2 and recommend screening in one year or give it a BI-RADS: 0 and recommend additional imaging.**

No images will be shown. Instead, we will give you additional information from an AI system and ask whether you would now recommend a recall and your confidence in that decision.

We will then present you with different AI results, but you should **visualize the same hypothetical mammogram throughout the entire study**. As you answer each question, do not imagine a new case. Base your responses solely on the new AI information provided in each scenario.

For this study, **assume a typical screening population** with a breast cancer prevalence of approximately 5-6 cancers in every 1,000 mammograms. Also, **assume that a typical mammography AI system** is being used with ROC (AUC) of 0.83. The AI system assigns a risk score for each patient from 0.00 to 1.00. **Higher scores indicate a greater likelihood of cancer, but the score is not an actual percentage**.

**In this clinical scenario, assume that your final BI-RADS assessment and the AI risk score would be visible to the patient on their patient portal.**

Please answer the following questions to the best of your ability.

#### Session 1 Vignette (Example)

You are reading a screening mammogram and are on the fence about whether to give it a BI-RADS: 1/2 and recommend screening in one year or give it a BI-RADS: 0 and recommend additional imaging.

You then review the exam’s AI score, which is **0.40** (possible range of 0.00-1.00, with higher scores indicating greater likelihood of cancer)

#### Session 2 Vignette (Example)

You are reading a screening mammogram and are on the fence about whether to give it a BI-RADS: 1/2 and recommend screening in one year or give it a BI-RADS: 0 and recommend additional imaging.

You then review the exam’s AI score, which is **0.40** (possible range of 0.00-1.00, with higher scores indicating greater likelihood of cancer)

**For an AI score greater than or equal to 0.40**: 97 in every 100 cases will be negative for cancer (and 3 in every 100 cases will be positive for cancer).

**For an AI score less than 0.40**: 3 in every 1,000 cases will be positive for cancer (and 997 in every 1,000 cases will be negative for cancer)

#### Questions

Given an AI score of **0.40**, would you recommend recall for additional imaging evaluation:

Yes / No

**Confidence** in your decision (0% none to 100% complete)

0% to 100% (slider)

## References

1. Haug CJ, Drazen JM. Artificial Intelligence and Machine Learning in Clinical Medicine, 2023. N Engl J Med. Massachusetts Medical Society; 2023;388(13):1201–1208. doi: 10.1056/NEJMra2302038.

2. Reddy S. Explainability and artificial intelligence in medicine. Lancet Digit Health. Elsevier; 2022;4(4):e214–e215. doi: 10.1016/S2589-7500(22)00029-2.

3. Li MD, Little BP, Alkasab TK, et al. Multi-Radiologist User Study for Artificial Intelligence-Guided Grading of COVID-19 Lung Disease Severity on Chest Radiographs. Acad Radiol. 2021;28(4):572–576. doi: 10.1016/j.acra.2021.01.016.

4. Lessmann N, Sánchez CI, Beenen L, et al. Automated Assessment of COVID-19 Reporting and Data System and Chest CT Severity Scores in Patients Suspected of Having COVID-19 Using Artificial Intelligence. Radiology. 2021;298(1):E18–E28. doi: 10.1148/radiol.2020202439.

5. van Assen M, Zandehshahvar M, Maleki H, et al. COVID-19 pneumonia chest radiographic severity score: variability assessment among experienced and in-training radiologists and creation of a multireader composite score database for artificial intelligence algorithm development. Br J Radiol. 2022;95(1134):20211028. doi: 10.1259/bjr.20211028.

6. Pacilè S, Lopez J, Chone P, Bertinotti T, Grouin JM, Fillard P. Improving Breast Cancer Detection Accuracy of Mammography with the Concurrent Use of an Artificial Intelligence Tool. Radiol Artif Intell. 2020;2(6):e190208. doi: 10.1148/ryai.2020190208.

7. Ahn JS, Ebrahimian S, McDermott S, et al. Association of Artificial Intelligence–Aided Chest Radiograph Interpretation With Reader Performance and Efficiency. JAMA Netw Open. 2022;5(8):e2229289. doi: 10.1001/jamanetworkopen.2022.29289.

8. Bernstein MH, van Assen M, Bruno MA, Krupinski EA, De Cecco C, Baird GL. Is a score enough? Pitfalls and solutions for AI severity scores. Eur Radiol Exp. 2025;9(1):67. doi: 10.1186/s41747-025-00603-z.

9. Van Calster B, Steyerberg EW, Wynants L, van Smeden M. There is no such thing as a validated prediction model. BMC Med. 2023;21(1):70. doi: 10.1186/s12916-023-02779-w.

10. Baird GL. The Importance of Quantifying Prediction in Real-World Settings. Am J Roentgenol. American Roentgen Ray Society; 2025;225(5):e2534011. doi: 10.2214/AJR.25.34011.

11. Yala A, Mikhael PG, Strand F, et al. Toward robust mammography-based models for breast cancer risk. Sci Transl Med. 2021;13(578):eaba4373. doi: 10.1126/scitranslmed.aba4373.

12. Kahneman D, Tversky A. Subjective probability: A judgment of representativeness. Cognit Psychol. 1972;3(3):430–454. doi: 10.1016/0010-0285(72)90016-3.

13. Song EC, Bernstein MH, Lay PS, et al. Accessing AI mammography reports impacts patient follow-up behaviors: the unintended consequences of including AI in patient portals. Npj Digit Med. Nature Publishing Group; 2025;8(1):761. doi: 10.1038/s41746-025-02201-0.

